# Investigating the effect of an online enhanced care program on the emotional and physical wellbeing of patients discharged from hospital with acute decompensated heart failure: Enhanced care program for heart failure

**DOI:** 10.1101/2025.03.05.25323198

**Authors:** Caroline Kuhne, Hannah Wetzler, Kristy Fakes, Christopher Oldmeadow, Andrew Boyle

## Abstract

**Background:** The Enhanced HF Care study is a prospective randomised open blinded endpoint (PROBE) design trial comparing the effectiveness of a digital health intervention –Enhanced HF Care -- to usual care, in increasing health related quality of life for patients hospitalised with acute decompensated heart failure (ADHF). The study aims to evaluate effectiveness in (i) improving emotional and physical wellbeing, and (ii) decreasing healthcare utilisation. This statistical analysis plan outlines the pre-specified statistical principles and procedures for analysing data from the trial, which has been updated from as described in the trial protocol to reflect lower than anticipated recruitment rates.

**Methods:** The co-primary outcomes are emotional and physical wellbeing measured using the Minnesota Living with Heart Failure Questionnaire (MLHFQ) domains at 6-months post-recruitment. Secondary outcomes include MLHFQ emotional and physical well-being at 1-month post-recruitment, unplanned hospital readmissions, and emergency department presentations. Statisticians conducting analyses are blind to treatment allocation.

**Analysis:** Bayesian hierarchical mixed effects models will be used to estimate treatment effects for all outcomes, with uninformative prior distributions specified for effect parameter and half-Cauchy priors on the random effect standard deviation and the within person standard deviation. For each outcome, the analysis will present mean differences between treatment groups with 95% credible intervals (highest posterior density) and posterior probabilities of treatment effect.

All randomised participants will be analysed according to their assigned treatment group following an intention-to-treat framework, excluding only those who withdraw consent for data use. Intercurrent events will be handled using the estimands framework. For MLHFQ outcomes, a composite strategy will be employed where participants who die will be assigned the maximum value reflecting worst possible quality of life. Healthcare utilisation outcomes (unplanned hospital readmissions and emergency department presentations) will be analysed using a ‘while alive’ strategy, with follow-up extending until either death or 6 months post-randomisation, whichever occurs first.

For handling missing data, we will use multiple imputation by chained equations (MICE) under a missing at random assumption when participants drop out. For completed MLHFQ subscales specifically, if only one item is missing within a subscale, we will use mean imputation for that item. However, if more than one item is missing form a subscale, the total score will be set to missing and MICE will be used to impute the total subscale score.

Additional analyses will estimate the posterior predictive probability of trial success had the target sample size been achieved.

**Conclusion:** Publication of this statistical analysis plan prior to data analysis ensures transparency in the analytical approach and minimises potential bias in the interpretation and reporting of trial results, particularly given the adaptation to Bayesian methods necessitated by lower-than-expected recruitment.

**Statement of Compliance:** This document is a statistical analysis plan for the aforementioned clinical trial. This trial will be conducted in compliance with all stipulation of this approved protocol, the conditions of the ethics committee approval, the NHMRC National Statement on Ethical Conduct in Human Research (2007), and the Note for Guidance on Good Clinical Practice (CPMP/ICH-135/95).

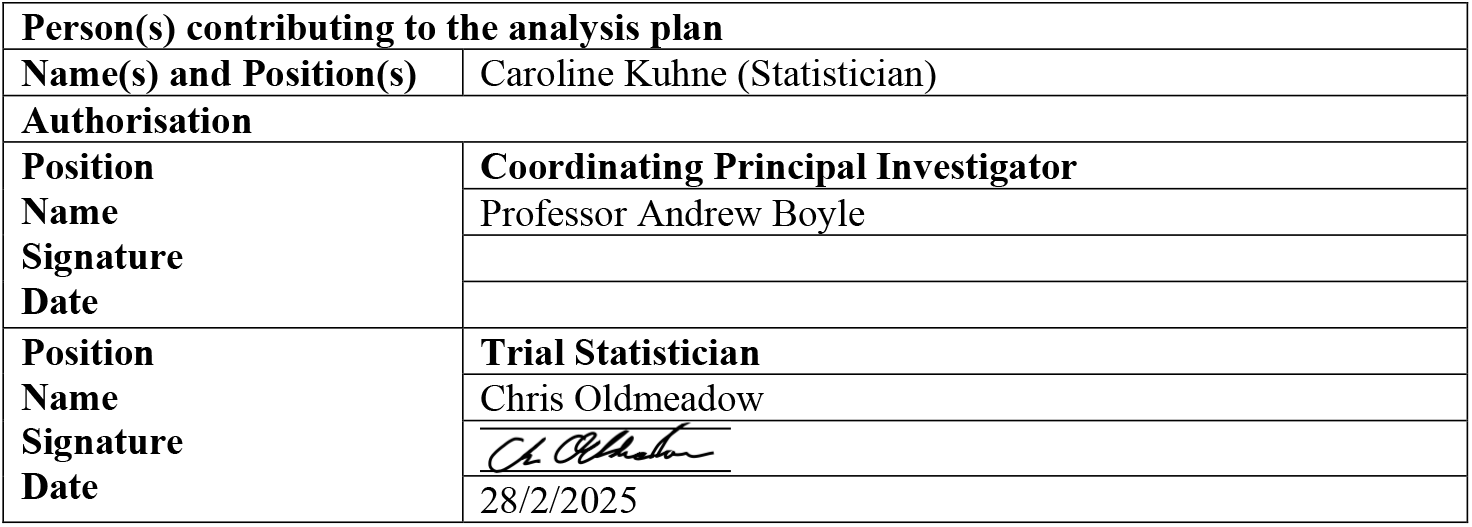

|  |  |
| --- | --- |
| **Person(s) contributing to the analysis plan** |  |
| **Name(s) and Position(s)** | Caroline Kuhne (Statistician) |
| **Authorisation** |  |
| **Position** | **Coordinating Principal Investigator** |
| **Name** | Professor Andrew Boyle |
| **Signature** |  |
| **Date** |  |
| **Position** | **Trial Statistician** |
| **Name** | Chris Oldmeadow |
| **Signature** |  |
| **Date** | 28/2/2025 |

**Administration information:** **Trial registration**

This trial was prospectively registered via the Australian New Zealand Clinical Trials Registry on the 4^th^ October 2022 (Reference: ACTRN12622001289707).

## 2. Introduction

### 2.1. Background and rationale

Heart failure (HF) affects over 64 million people globally and represents a significant public health burden. Depression is highly prevalent in HF patients and is associated with increased hospitalisations, reduced health functioning, and higher mortality rates. Given the interaction between emotional and physical wellbeing in disease progression, there is a critical need for enhanced emotional support and information for HF patients. Digital health interventions show promise in helping HF patients manage both their emotional and physical wellbeing, though evidence from existing trials is mixed, and studies have been limited by small sample sizes.

The Enhanced HF Care program, a digital health intervention, seeks to improve care coordination with a proactive approach to monitoring and managing emotional and physical wellbeing. The proposed program is designed to achieve this through: (i) recognising the interaction between emotional and physical wellbeing; (ii) harnessing existing community resources, such as GPs; (iii) supporting patient empowerment through offering education and self-help strategies; (iv) providing a mechanism for tailored self-help and stepwise care; (v) using clinically derived algorithms to offer care recommendations; and (vi) increasing the probability of a care partnership between hospital providers, community health, the patient and their support persons.

#### Purpose of the Statistical Analysis Plan

This document describes the statistical methods and procedures to be used in analysing data from the Enhanced HF Care trial which has been updated to reflect lower than anticipated recruitment rates. It has been developed prior to data analysis to ensure analytical transparency and minimise potential reporting bias.

## 3. Study Methods

### 3.1. Trial Design

This study is a prospective randomised open blinded endpoint (PROBE) design trial with a 1-month and 6-month follow-up. The trial has two-arms:

1. **Intervention:** Online enhanced community care program for Heart Failure: ‘Enhanced HF Care’, plus usual care.
2. **Control:** Usual care alone

Treatment arms were randomly allocated in a 1:1 ratio. While participants and care providers are not blinded to treatment allocation, outcome assessors and analysts are blinded to allocation.

Figure 1^1^ presents a CONSORT diagram with an overview of the trial design and interventions (created prior to study commencement). Figure 2 presents the CONSORT diagram showing participant flow through the study, including screening, randomisation, and follow-up.

**Figure 1:**
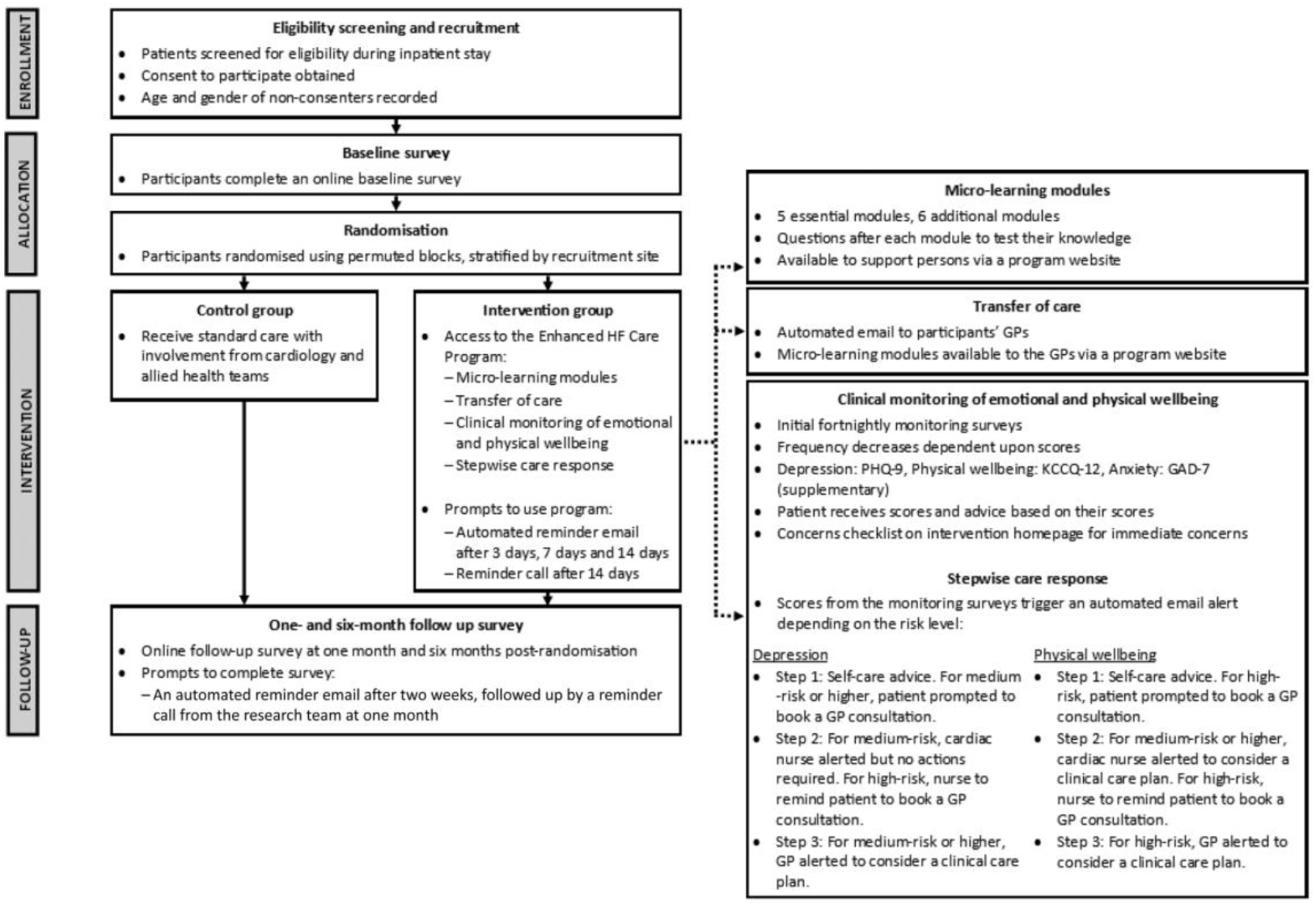
CONSORT diagram of participant flow through the study (prior to study commencement). Reprinted from Fakes K, Hobden B, Zwar N, Collins N, Oldmeadow C, Paolucci F, Davies A, Fernando I, McGee M, Williams T, Robson C. Investigating the effect of an online enhanced care program on the emotional and physical wellbeing of patients discharged from hospital with acute decompensated heart failure: Study protocol for a randomised controlled trial: Enhanced care program for heart failure. Digital Health. 2024 May; 10:20552076241256503.

**Figure 2:**
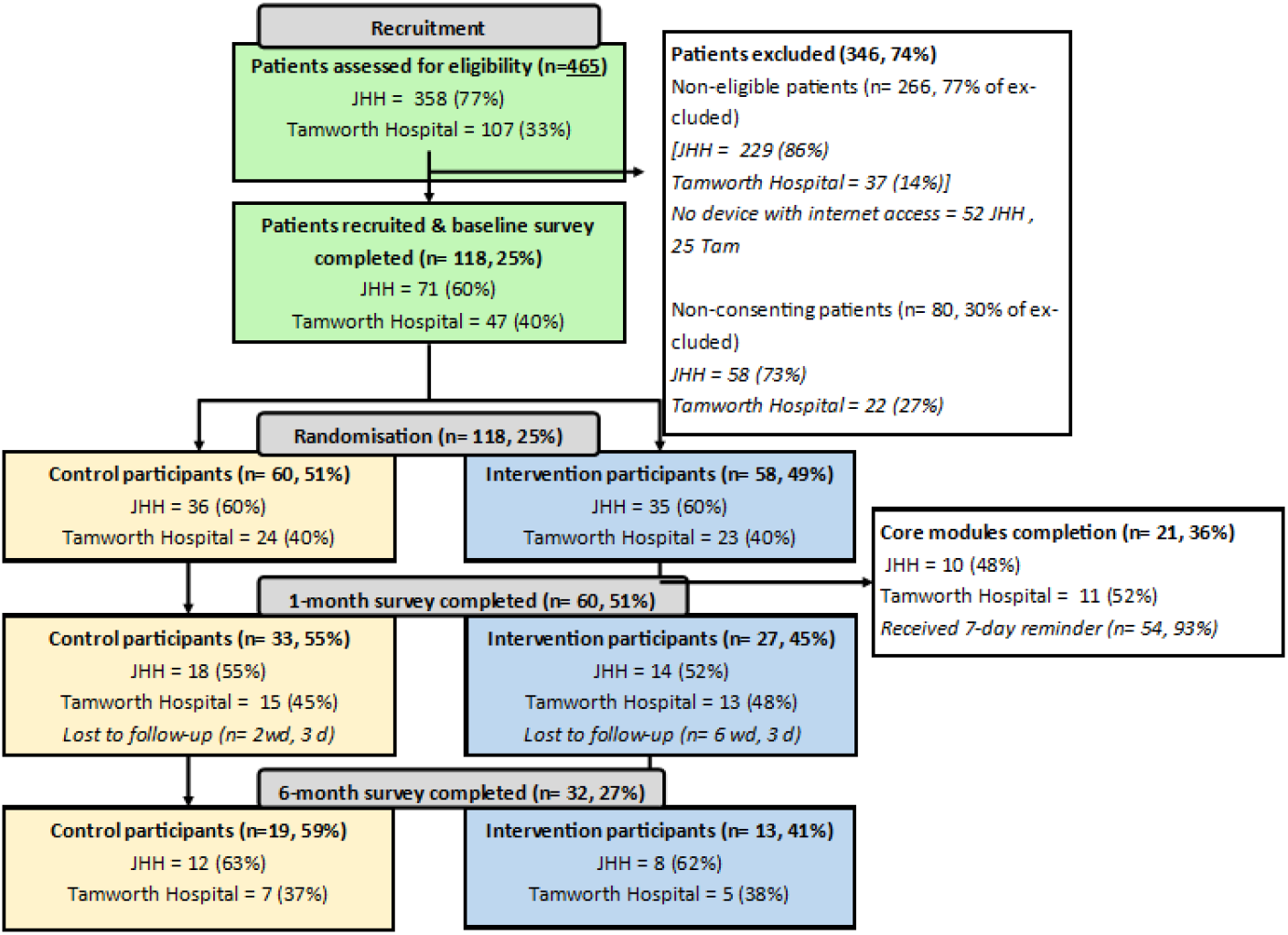
CONSORT diagram of participant flow through the study (interim study numbers).

### 3.2. Intervention Groups

#### 3.2.1. Intervention: ‘Enhanced HF Care’, plus usual care

Participants randomised to Condition 1 will receive usual care, plus the Enhanced HF Care program. See Figure 1 for Intervention Flowchart. For full detail on the Enhanced HF Care program, see original protocol^1^.

#### 3.2.2. Control: Usual care group

Participants randomised to Condition 2 will receive standard care with involvement from cardiology and allied health teams which may differ by site. This includes an initial call within 2 weeks post-hospital discharge to educate and assess patients in regard to their understanding of their condition and treatment, a medication reconciliation and symptom management.

Patients are recommended to have a follow-up appointment with their GP 1-week post-discharge and encouraged to formulate an action plan, and an appointment 1 month later with their cardiologist. Further support is offered in relation to exercise, lifestyle modification and palliative care depending on the patient’s condition and prognosis. One- and 6-monthly data collection will obtain post-discharge care details including the use of health services.

### 3.3. Randomisation

Randomisation to either 1) intervention, or 2) usual care, was implemented in REDCap following the completion of the baseline survey, using a schedule developed by an independent statistician. The schedule used randomly selected block sizes of two and four, stratified by recruitment site, with a randomisation ratio of 1:1. This approach ensures balance in treatment allocation both overall and within sites. Due to the nature of the intervention, it was not possible to blind trial participants or cardiac nurses to the randomisation allocation.

The randomisation allocation was known to the participant, cardiac nurses, or researchers involved in supporting the trial. However, outcome assessors and data analysts are blinded to allocation.

### 3.4. Trial Population

#### 3.4.1. Eligibility

Patients will be eligible to participate if the following apply:

- Hospitalised with ADHF (including all ejection fraction classifications) being discharged from participating hospitals to home
- Provide written, informed consent to participate in the study.
- Aged 18 years or older
- English speaking; with access to, and ability to utilise, a device with internet
- Able to provide informed consent
- Well enough to be approached about the study (nurse’s judgement).

#### 3.4.2. Exclusion Criteria

Patients will be ineligible to take part in the study if the following apply:

- Without access to a device with internet
- Unable to provide written informed consent.
- Non-verbal.

### 3.5. Sample Size

The original study design targeted a sample size of 570 participants (285 per group). This sample size was calculated based on a previous study of approximately 1,200 patients that established sensitivity and minimum clinically important differences in the MLHFQ34. Using the reported standard deviations of 7.2 points (emotional domain) and 9.5 points (physical domain), and accounting for a 0.5 correlation between baseline and follow-up measurements, this sample size would provide 80% power to detect clinically significant differences of 1.75 points in the MLHFQ emotional domain and 3.5 points in the physical domain at 6 months, while maintaining a type 1 error rate of 5%. The calculation included an adjustment for an anticipated 30% drop-out rate.

As of 4.12.2024, only 118 patients have been recruited (32 at 6-month follow-up), falling substantially short of the target sample size. This lower-than-anticipated recruitment has prompted a revision of the statistical analysis plan, shifting from the originally planned frequentist approach to a Bayesian framework. This Bayesian approach allows for more nuanced interpretation of the available evidence given the achieved sample size, without requiring the same strict sample size requirements as frequentist hypothesis testing.

## 4. Objectives, Endpoints and Estimands

### 4.1 Primary objective

The primary objective of the study is to estimate the effect of treatment (Enhanced HF Care) compared to usual care in improving self-reported emotional and physical wellbeing, at 6-months post-recruitment among patients hospitalised with acute decompensated heart failure (ADHF).

The co-primary endpoints are the physical and emotional domains of the Minnesota Living with Heart Failure Questionnaire (MLHFQ). The MLHFQ assesses the impact of heart failure on health-related quality of life. All items are answered on six-point Likert scales from 0 (none) to 5 (very much), with higher scores reflecting a worse health related quality of life. It has strong psychometric properties and is sensitive to change.

The physical domain of the MLHFQ has 8 items, with scores ranging from 0-40. The emotional domain of the MLHFQ has 5 items, with scores ranging from 0-25.

#### 4.1.1. Primary objective endpoints and estimands

| Endpoint | Population | Intercurrent events | Summary measure |
| --- | --- | --- | --- |
| The emotional domain of the MLHFQ at 6 months post randomisation. | The intention-to-treat (ITT) population, defined as all randomised participants excluding participants who have withdrawn their data, with their data analysed according to the experimental group to which they were randomised to. | 1) Deaths: Some patients might die before 6 months, so their quality-of-life scores are not defined past the point they die. We will use the ‘**Composite Strategy**’ assigning a maximum value (reflecting worst possible QOL) to those who die before 6 months.<br>2) Inadequate compliance:<br>**Treatment policy strategy.** Analysed according to the experimental group to which they were randomised to, regardless of compliance. | The mean difference between intervention and control in emotional domain scores at 6 months. |
| The physical domain of the MLHFQ at 6 | As above | As above | The mean difference between intervention |
| months post randomisation. |  |  | and control in emotional domain scores at 6 months. |

### 4.2 Secondary objectives

The secondary objective is to estimate among patients hospitalised with acute decompensated heart failure (ADHF):

1. The effect of Enhanced HF Care compared to usual care in improving self-reported emotional and physical wellbeing, at 1-month post-randomisation. For this objective, scores on the physical and emotional domains of the MLHFQ at 1-month post randomisation will be used.
2. The effect of Enhanced HF Care compared to usual care in reducing healthcare utilisation at 1-month and 6-months post randomisation.

This objective assesses whether Enhanced HF Care reduces healthcare utilisation compared to usual care. Medical records will be used to determine the number of unplanned hospital readmissions, emergency department presentations, and the length of hospital stay.

Additionally, self-reported data on the use of primary health, allied health, and community services will be collected.

#### 4.2.1 Secondary objective endpoints and estimands

4.2.1. Secondary objective endpoints and estimands
| Endpoint | Population | Intercurrent events | Summary measure |
| --- | --- | --- | --- |
| The emotional domain of the MLHFQ at 1 month post randomisation. | The intention-to-treat (ITT) population, defined as all randomised participants excluding participants who have withdrawn their data, with their data analysed according to the experimental group to which they were randomised to. | 1) Deaths: We will use the ‘**Composite Strategy**’ assigning a maximum value (reflecting worst possible QOL) to those who die before 6 months<br>2) Inadequate compliance: We will use the **Treatment policy strategy**. analysed according to the experimental group to which they were randomised to, regardless of compliance | The mean difference between intervention and control in emotional domain scores at 1 month |
| The physical domain of the MLHFQ at 1 months post randomisation. | As above | As above | The mean difference between intervention and control in physical domain scores at 1 month |
| The number of unplanned hospital readmissions from randomisation to 1- and 6- months post-randomisation. | As above | 1) Deaths: We will use the ‘**While alive**’ strategy, restricting the follow-up to be up until the point of death or end of 6 months (whichever occurs sooner)<br>2) Inadequate compliance: We will use the **Treatment policy strategy**.<br>Analysed according to the experimental group to which they were randomised to, regardless of compliance | The ratio of the cumulative incident rate of unplanned hospital readmissions between intervention and control at 1 month and 6 months |
| The number of hospital emergency department presentations from randomisation to 1- and 6-months post-recruitment. | As above | As above | The ratio of the cumulative incident rate of ED presentations between intervention and control at 1 month and 6 months |
| Total self-reported visits to or instances of care received from primary health care providers at 1-month and 6-months post-randomisation. | As above | As above | The ratio of the cumulative incident rate of ED presentations between intervention and control at 1 month and 6 months |
| Total self-reported visits to or instances of care received from allied health providers at 1-month and 6-months post-randomisation. | As above | As above | The ratio of the cumulative incident rate of ED presentations between intervention and control at 1 month and 6 months |
| Total self-reported visits to or instances of care received from community service providers at 1-month and 6-months post-randomisation. | As above | As above | The ratio of the cumulative incident rate of ED presentations between intervention and control at 1 month and 6 months |
| Length of Hospital Stay | As above | As above | The mean difference between intervention and control in physical domain scores at 1 month |

## 5. Statistical methods

### 5.1 Changes to the Statistical considerations section of the trial protocol

#### 5.1.1 Treatment of missing data

The protocol stated that we would use a method that does not rely on the assumption the data are missing at random (e.g. pattern mixture models). Since then, we have used the estimands framework to clarify the treatment of intercurrent events (using the composite strategy -for the QOL co-primary outcomes, assigning a maximum value (reflecting worst possible QOL) to those who die. This strategy most aligned with the research question and objectives. Missing data due to loss-to-follow-up or withdrawal will be treated as missing at random.

#### 5.1.2 Change to Bayesian Framework

The statistical methods section in the protocol was stated as a linear mixed model within a frequentist inferential framework. Due to the ongoing persistent recruitment problems outside of the research teams control, we have changed to a Bayesian inferential framework. This will allow more meaningful and nuanced interpretation of the trial results for an otherwise **underpowered** study.

### 5.2. Analysis Timeline

#### 5.2.1. Statistical interim analyses and stopping guidance

Interim analyses are planned for February 2025, where the approximately expected recruitment rate at this time will be 50%. The results of this interim analysis will not form a stopping rule, as the purpose is for reporting and dissemination of results to date.

#### 5.2.2. Timing of final analysis

The primary analysis will occur in December 2025.

#### 5.2.3. Timing outcome assessments

Data will be collected from participants at three time-points: (i) at baseline following consent in an online REDCap survey; (ii) at 1-month follow up in an online REDCap survey and, (iii) 6-months follow up in an online REDCap Survey. See Table 1 for a schedule of outcome assessments.

**Table 1.** Schedule of Outcome Assessments.

| **Timepoint** | **DESCRIPTION** |
| --- | --- |
| **Baseline (in hospital prior to discharge)** | • Physical domain of the Minnesota Living with Heart Failure Questionnaire (MLHFQ).<br>• Emotional domain of the MLHFQ. |
| **1-month post randomisation** | • Physical domain of the Minnesota Living with Heart Failure Questionnaire (MLHFQ).<br>• Emotional domain of the MLHFQ.<br>• Unplanned Hospital readmissions<br>• Hospital emergency department presentations<br>• Length of hospital stay<br>• Total visits/care received from primary health care providers<br>• Total visits/care received from allied health care provider<br>• Total visits/care received from community service providers |
| **6-months post randomisation** | • Physical domain of the Minnesota Living with Heart Failure Questionnaire (MLHFQ).<br>• Emotional domain of the MLHFQ.<br>• Unplanned Hospital readmissions<br>• Hospital emergency department presentations<br>• Length of hospital stay<br>• Total visits/care received from primary health care providers<br>• Total visits/care received from allied health care provider<br>• Total visits/care received from community service providers |

### 5.3. Endpoint Definitions

Co-primary endpoints will be measured at baseline, and at 1- and 6-month follow-up times. While the co-primary endpoints are measured at all three time points, the primary outcome is defined at 6 months post-randomisation. The 1-month follow-up assessments of these endpoints are considered secondary outcomes. Additional ssecondary outcomes will be measured with data from the medical records and via patient self-report survey at 1- and 6-months follow-up times. An overview of the outcomes to be assessed are shown in Table 2.

**Table 2.** Description of outcomes to be collected and analysed.

| **Endpoint** |  |  | **DESCRIPTION** |
| --- | --- | --- | --- |
| **Primary** |  | Minnesota Living with Heart Failure Questionnaire (MLHFQ) | The co-primary outcomes are the physical and emotional domains of the Minnesota Living with Heart Failure Questionnaire (MLHFQ). The MLHFQ assesses the impact of heart failure on health-related quality of life. All items are answered on six-point Likert scales from 0 (none) to 5 (very much), with higher scores reflecting a worse health related quality of life. It has strong psychometric properties and is sensitive to change. |
|  | **1** | Physical domain of the MLHFQ – 6 months follow-up | The physical domain of the MLHFQ has 8 items, with scores ranging from 0-40. |
|  | **2** | Emotional domain of the MLHFQ – 6 months follow-up | The emotional domain of the MLHFQ has 5 items, with scores ranging from 0–25. |
| **Secondary** | **1** | Physical domain of the MLHFQ – 1 month follow-up | As above in primary outcome. |
|  | **2** | Emotional domain of the MLHFQ – 1 month follow-up | As above in primary outcome. |
|  | **3** | Unplanned Hospital readmissions – 1 and 6 months follow-up | The number of unplanned hospital readmissions will be obtained from medical records. |
|  | **4** | Hospital emergency department | The number of hospital emergency department presentations will be obtained from medical records. |

| Endpoint |  |  | DESCRIPTION |
| --- | --- | --- | --- |
|  |  | presentations – 1 and 6 months follow-up |  |
|  | 5 | Length of Hospital Stay | Length of Hospital Stay will be obtained from medical records. |
|  | 6 | Total self-reported visits to or instances of care received from primary health care providers at 1-month and 6-months post-randomisation. | This will be obtained from self-report survey data about primary health care service usage. |
|  | 7 | Total self-reported visits to or instances of care received from allied health providers at 1-month and 6-months post-randomisation. | This will be obtained from self-report survey data about allied health care service usage. |
|  | 8 | Total self-reported visits to or instances of care received from community service providers at 1-month and 6-months post-randomisation. | This will be obtained from self-report survey data about community care service usage |

### 5.4. Analysis populations

The primary analysis modelling will be conducted as intention-to-treat (ITT), defined as all randomised participants excluding participants who have withdrawn their data, with their data analysed according to the experimental group to which they were randomised to.

### 5.5. Analysis Methods

#### 5.5.1. Descriptive statistics and Presentation of Results

Descriptive statistics will be used to summarise the baseline demographic and clinical characteristics overall and for intervention and control groups in the intention to treat population. Counts and percentages for categorical data, and means, standard deviations (or medians, interquartile ranges) for continuous data. Example templates for summary tables are provided in Appendix 6.1, which illustrate the planned format for presenting baseline characteristics (Table A1), primary outcomes (Table A2), and secondary outcomes (Table A3).

#### 5.5.2. General Statistical methodology

Statistical inference for assessing differences between groups for all outcome measures will be within a Bayesian framework. This involves, for each outcome, specifying a likelihood function and prior distributions, which together form the posterior distribution for the parameters of interest. This approach allows for more nuanced interpretation of the evidence given the achieved sample size.

#### 5.5.3. Prior Specification

We will use uninformative prior distributions for all treatment effect parameters and half-Cauchy priors for the standard deviation of the random effects and the within-person standard deviation. These are outlines in more detail below.

#### 5.5.4. Samples from the posterior distribution

Samples from the posterior distribution will be obtained using the No U-Turn Sampler (NUTS), as implemented in the brms R package^2^. All models will be run with 5,000 burn-ins and 7,000 iterations, using the default settings of 4 chains. No thinning will be applied. A seed will be set for reproducibility.

#### 5.5.5. Assessment of convergence

We will evaluate convergence through:

- Visual inspection of trace plots and posterior distributions
- Gelman-Rubin convergence statistic (Rhat ≤1.1 indicating convergence)
- Effective sample size (ESS >400 for stable estimates)

If convergence is not achieved, we will initially attempt to increase the number of iterations, and if issues remain, we will vary the thinning rate, then then try stronger [informative] prior distributions.

#### 5.5.6. Assessment of fit

To assess model fit, the posterior predicted values of the outcome will be compared to the observed data. If models indicate poor fit, stronger prior distributions or alternate model distributions will be explored.

#### 5.5.7. Summaries of the posterior distribution

##### Posterior mean

The mean of the converged posterior distribution for the parameter reflecting the between group differences (either absolute or relative) will be presented as the point estimate to 2 decimal places.

##### Intervals

95% credible intervals will be calculated using the highest posterior density (HPD) method and presented to 2 decimal places.

##### Probability of direction

The one-sided probability that the difference between treatment and control is greater than zero will be presented (representing a beneficial treatment effect). This is calculated as the proportion of posterior samples that are of sign that is favorable to the intervention (either +’ve or –’ve)

### 5.6. Statistical models

#### 5.6.1. Primary outcome

The co-primary outcome variables – self-reported emotional and physical well-being at 6-months post baseline--will be assessed in the ITT population using a Bayesian hierarchical mixed effects linear regression. A random intercept for participant ID will be included to account for serial correlations from the repeated measures on the same individual. The random intercept will be modelled as normally distributed with mean 0, with a half-Cauchy prior on the standard deviation. The residual variation (within-person standard deviation) will also be assigned a half-Cauchy prior. Fixed effects for treatment group, categorical time, baseline outcome value, hospital location, and the interaction of treatment group and time, will have non-informative priors for the parameters respective parameters (as mean difference for continuous outcomes).

The hierarchical model is specified below:

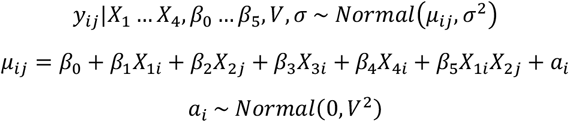

*i* = participant

*j* = timepoint

*X*_1*i*_ = Treatment group for participant i

*X*_2*j*_ = Timepoint j

*X*_3*i*_ = Baseline outcome for participant i

*X*_4*i*_ = Hospital Location (Stratification Variable) for participant i.

Priors

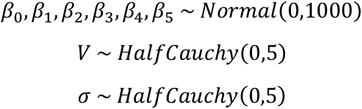

#### 5.6.2. Secondary outcomes

Continuous secondary outcomes will be modelled as linear with specifications as outlined in the analysis of primary outcomes. Count data (e.g., hospital readmissions) will be analysed as a negative binomial regression with an offset term for follow up time to account for the “while alive strategy”. In the event of excessive zeros, a zero-inflated negative binomial regression will be used.

#### 5.6.3. Posterior Predictive Analysis To estimate the probability of trial success if target recruitment (N=570) had been achieved

In additional secondary analyses, we will estimate the posterior predictive probability of a successful trial had the trial recruited the target sample. To achieve this, 1000 draws will be taken from the posterior distributions of the parameters of the co-primary 6-month endpoint models. For each draw data for additional participants up to the target sample size 570 will be simulated. The existing data will be incorporated by treating it as known and only simulating the additional participants needed to reach the target sample size. The randomisation ratio of 1:1 will be maintained.

A success criterion of .9 posterior probability of treatment effect is defined for each of the co-primary endpoints. For each of the simulated complete data sets we will repeat the same Bayesian analysis and determine if it meets the success criteria. We then calculate the probability of success as the proportion of simulated datasets that meets the success criteria to give the posterior predictive probability of trial success for each primary outcome, if continued to target N of 570.

### 5.7. Missing Data

Missing data due to death:

The estimands framework will be used to handle intercurrent events. For MLHFQ, a composite strategy will be employed where participants who die will be assigned the maximum value reflecting worst possible quality of life (QOL). For healthcare utilisation outcomes (unplanned hospital readmissions and emergency department presentations), a ‘while alive’ strategy will be used, with follow-up extending until either death or 6 months post-randomisation, whichever occurs first.

Other missing

Missing data due to participant drop out will be addressed using multiple imputation by chained equations (MICE) under a missing at random assumption.

For completed MLHFQ subscales specifically, if only one item is missing within a subscale, we will use mean imputation for that item. However, if more than one item is missing form a subscale, the total score will be set to missing and MICE will be used to impute the total subscale score.

### 5.8. Sensitivity analysis

A sensitivity analysis will be performed with the primary analysis run again with only complete cases to determine the robustness of the multiple imputations.

### 5.9. Statistical Software

The cleaning, coding, and analysis of the data sets will be completed using Statistical Analysis System (SAS) version 9.4 and R (R Foundation for Statistical Computing, Vienna, Austria. URL https://www.R-project.org/).

## Data Availability

The data from this study will not be openly available. Results will be published in peer-reviewed journals and presented at national and international conferences. Additionally, participating hospitals will receive a de-identified report of results specific to their recruited patients. Researchers interested in further details may contact the study authors for inquiries.

## 6. Appendix

The following tables provide example templates for the presentation of results.

**Table A1a:**
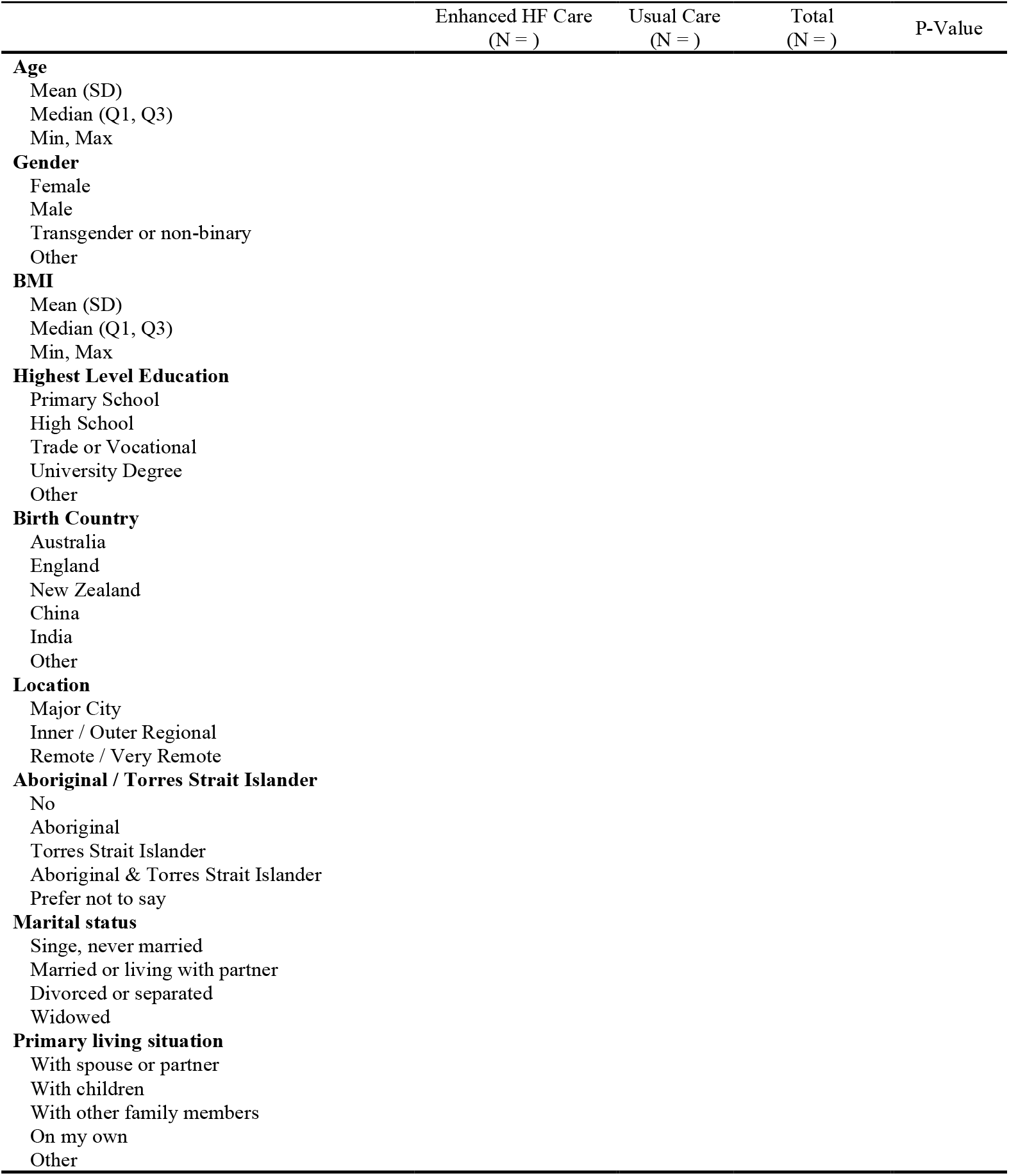
Basic Demographic by Treatment Group (ITT Population)

**Table A1b:**
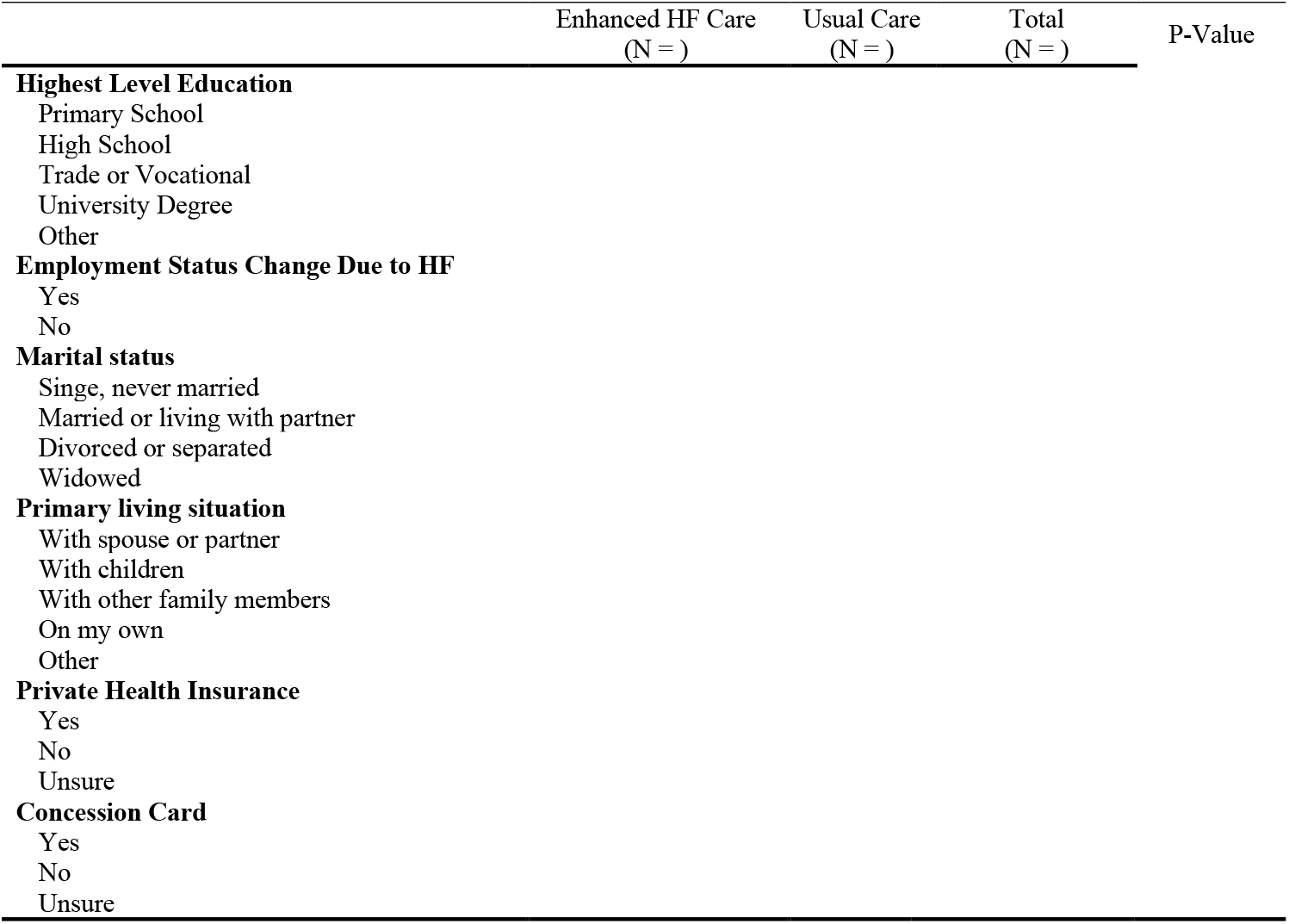
Socioeconomic status and living situation by treatment group (ITT Population)

**Table A1c:**
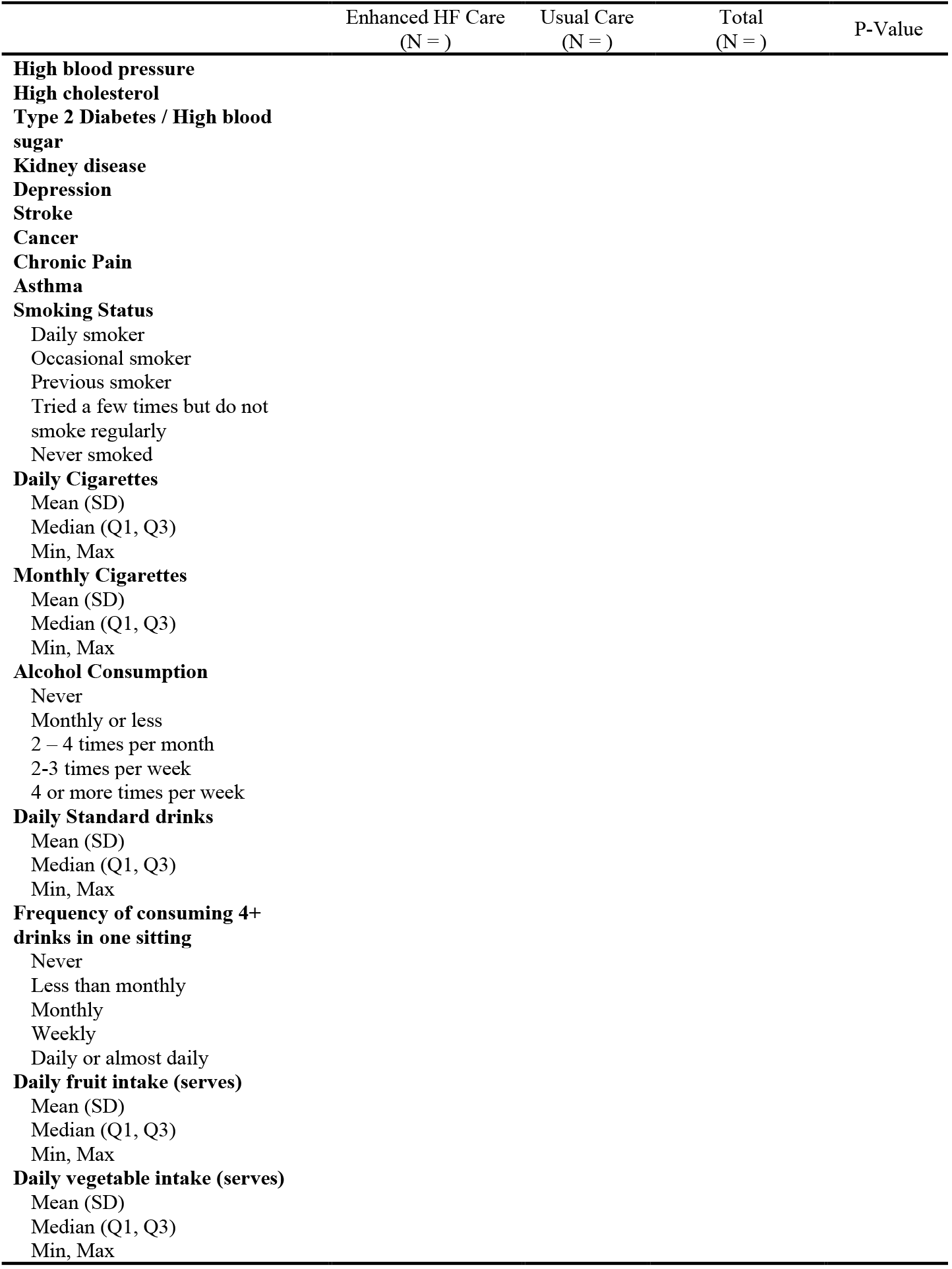
Health Conditions and risk factors by Treatment Group (ITT Population)

**Table A2:**
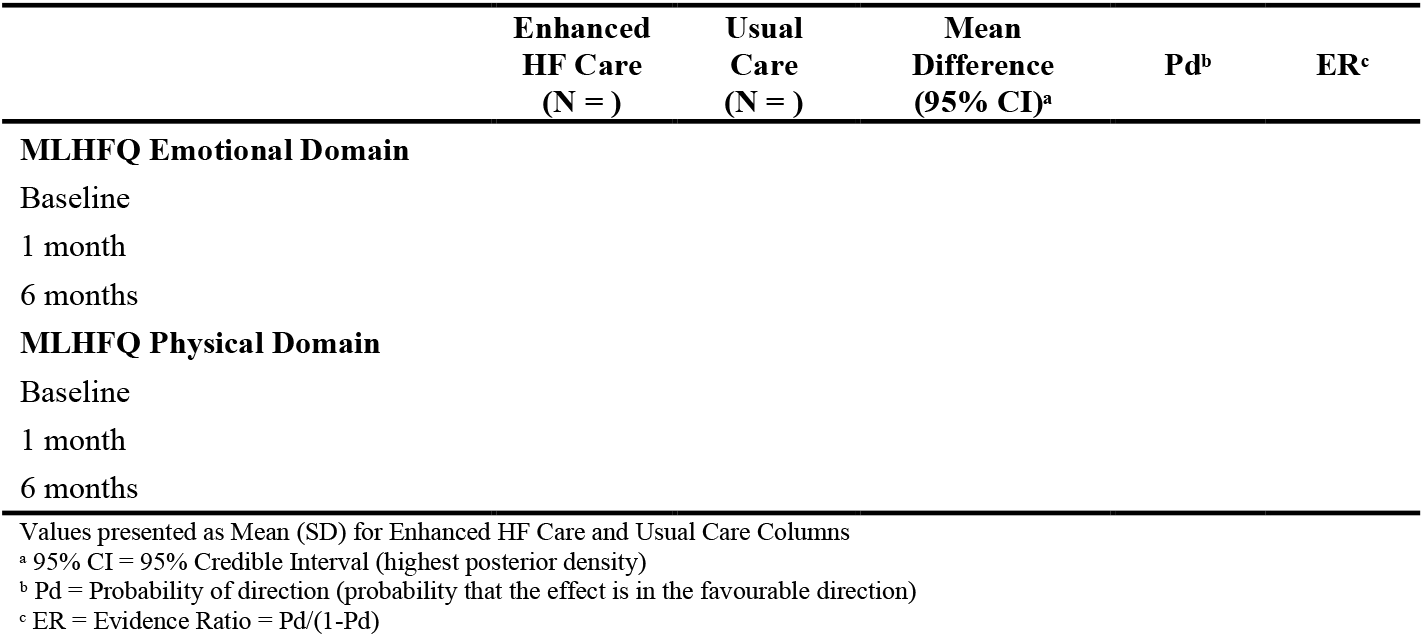
MLHFQ domain outcomes at 1-month (secondary outcome) and 6-months (primary outcome) by treatment arm.

**Table A3:**
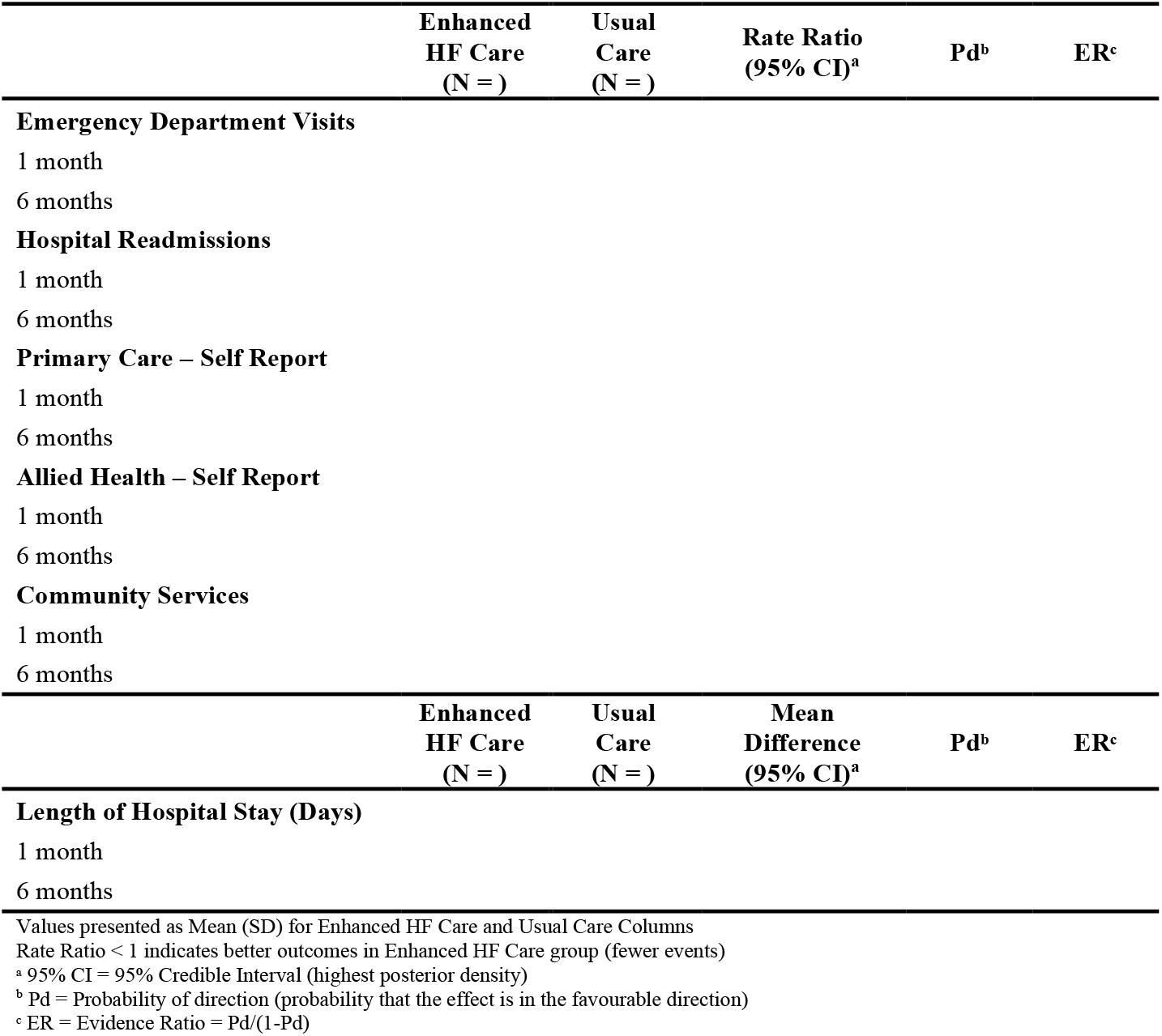
Secondary Outcomes of Healthcare Utilisation by Treatment Group and Timepoint.

